# Estimation of the global burden of autosomal recessive rare inborn errors of metabolism

**DOI:** 10.1101/2025.02.14.25322285

**Authors:** Soumyajit Mondal, Kalyan Goswami, Atanu Kumar Dutta

**Author notes:** Corresponding author Dr Atanu Kumar Dutta, Additional Professor, Department of Biochemistry, Second Floor; Academic Block I, All India Institute of Medical Sciences, Kalyani, Saguna, NH-34 Connector, Basantapur, Kalyani, West Bengal 741245, Contact number: 8967369455.

## Abstract

While many Rare Inborn Errors of Metabolism are treatable conditions their optimal diagnosis and treatment is a challenge for nations with low resources. Moreover, the population prevalence of these conditions is largely unknown. The availability of large genomic datasets brings the opportunity to estimate population carrier frequency of autosomal recessive IEMs. This would help to generate diseases burden statistics for better allocation of resources. In the current work we estimated the gene specific combined minor allele frequency of pathogenic variants from the gnomAD dataset for 235 genes associated with IEM phenotypes in OMIM. As per our estimation almost one third of the Global population is carrier for a pathogenic variant responsible for rare autosomal recessive inborn error of metabolism with the highest carrier frequency in the Ashkenazi Jews. Globally per thousand live births approximately five children are born with an ARIEM. European Finnish have the highest burden of nine out of 10,000 live births. With 25 million live births per year India is expected to have at least 8,025 newborns with an ARIEM. Since many of these diseases are treatable early newborn screening holds the key to ensure optimal management of these children.

## INTRODUCTION

With the completion of the Human Genome Project (Lander et al.,2001, Venter et al.,2001), significant advancements in genetic research were noticed, particularly in distinguishing benign genomic variations from those associated with monogenic and complex diseases (Tabor et al.,2014). Since then, large-scale genomic databases have been established, providing a comprehensive and precise view of human genetic diversity. These resources are crucial for determining allele frequencies, which are instrumental in pinpointing genetic factors underlying Mendelian disorders. Meanwhile, the advent of Next-generation sequencing (NGS) revolutionized medical genetics by generating vast amounts of variant genomic data for various diseases. However, most of these variants are often non-pathogenic, necessitating accurate differentiation between pathogenic mutations and remaining variations (Marian et al.,2020). This includes precise estimation of carrier frequencies (CF) for mutations linked to specific genetic disorders, which is essential for effective clinical diagnosis and management. The integration of these genomic insights will enhance our understanding of disease mechanisms and improve the accuracy of genetic diagnostics (Lewis & Vassos, 2020).

Inborn errors of metabolism (IEM) represent a group of genetic disorders characterized by defects in metabolic pathways, leading to the accumulation of toxic substances or the deficiency of essential metabolites. The clinical presentation of IEM can vary widely, with some conditions manifesting in infancy and others presenting later in life (Ferreira & Karnebeek, 2019). Though IEMs are individually rare but collectively common. A promising aspect is that many IEMs are amenable to treatment through the restoration of homeostasis in disrupted metabolic pathways. The complexity and heterogeneity of IEM necessitate a comprehensive understanding of their genetic underpinnings, as well as the development of effective screening and treatment strategies to mitigate their impact on affected individuals and families (Kruszka & Regier,2019).

IEMs pose a significant public health challenge in South Asia, the prevalence and spectrum of IEMs vary across different regions, influenced by factors such as consanguinity and genetic diversity. A study conducted in South India reported a prevalence of IEM at approximately 1 in 2497 newborns, although the true prevalence across the region remains largely undocumented (Krishnamurthy et al., 2020). The lack of comprehensive newborn screening programs exacerbates the situation, as many cases go undiagnosed until symptoms manifest, often leading to severe morbidity or mortality (Therrell et al., 2024). Furthermore, the high rates of consanguinity in South Asia increase the risk of recessive genetic disorders, further complicating the epidemiological landscape of IEM in this region (Vankwani, 2024).

It always remains a key question in human genetics to determine the proportion of people in a population who carry a single copy of a particular recessive genetic variant called as carrier frequency (CF) and the proportion of people in a population expected to have the disease called as disease prevalence (DP). This CF and DP are population specific and highly important for strategy building. For developing nations, it is way more challenging to allocate resources for rare diseases due to high prevalence of infectious and emerging lifestyle diseases. Estimation of the rare disease burden therefore would provide critical data for deciding the allocation of scarce resources.

In our current work, we retrieved population specific carrier frequencies of South Asian population for 1,72,963 variants in 221 AR IEM genes from gnomAD v2.1 dataset (https://gnomad.broadinstitute.org/). We classified these variants as pathogenic based on probable loss of function (plof) status, allele frequency of <0.005, ClinVar annotation, and InterVar annotation. We therefore narrowed down to 8,067 pathogenic variants in 221 AR IEM genes. We then computed combined minor allele frequencies. Considering the population follows Hardy-Weinberg equilibrium, we estimated carrier frequency and disease prevalence using it. We also compared all these data with each of six other major worldwide subpopulations. In the current work, we have also designed an analytical framework that enables researchers to utilize genetic data from multiple large-scale genomic databases.

## METHODOLOGY

### AR-IEM Gene Detection

The list of Rare Inborn Errors of Metabolism (ORPHA 68367) and corresponding genes were extracted from Orphanet and OMIM databases in February 2020.

### Creating AR-IEM Mutation Databases

First, we created a Structured Query Language (SQL) of all the DNA variants of all AR-IEM genes from the gnomAD version 2.1 dataset (https://gnomad.broadinstitute.org/). The DNA variants were stratified according to all the ethnic groups of the individuals carrying them as per the information given in the gnomAD version 2.1 database. Two separate SQL databases will also be created for all the DNA variants of all AR-IEM genes from ClinVar (https://www.ncbi.nlm.nih.gov/clinvar/) and InterVar (https://wintervar.wglab.org/) database respectively.

### Analysing methodology of the databases

To identify true pathogenic mutations, several filtering steps were be used (figure –). At first, variants from the gnomAD v2.1 database that were found only in one homozygous individual but not in any heterozygotes were eliminated as these data will likely represent nonreliable reads. Next, the remaining variants were divided into two subgroups: one with probable truncating variants [nonsense, frameshift, canonical ±1 or 2 splice sites, initiation codon, single exon or multiexon deletion; based on the American College of Medical Genetics classification for truncating variants (Richards et al.,2015)] and “other” variants.

We then validated the categorization information regarding the probable truncating variants as provided by the gnomAD. For this, stratification of the probable truncating variants in two groups were done using sequence quality (as per gnomAD data) and allele frequency of 0.005 as two parameters. Those with both high-quality sequencing and allele frequency of less than or equal to 0.005 were considered more likely to be pathogenic. On the other hand, the variants that either have low-quality sequencing or allele frequency > 0.005.

For the “other” variants group, they were searched in the SQL database generated from ClinVar and among those found in ClinVar with labelling of “likely pathogenic” or “pathogenic” will be taken as more likely to be pathogenic variants. Those variants that will not be there in ClinVar, will be searched in the other SQL database generated from InterVar previously. If any of them was found in InterVar to be interpreted as “likely pathogenic” or “pathogenic” they were considered more likely to be pathogenic.

Adding all these together we came to the total number of pathogenic AR-IEM variants. Based on these data, we calculate the Carrier Frequency (CF) value as detailed below.

### Calculation of Carrier Frequency (CF)

The gnomAD database presents the following data for each subpopulation: allele count (total number of a specific allele detected in a given subpopulation), allele number (total number of genotyped alleles at the genomic position of the variant considered), and homozygote count (total number of homozygous individuals for a specific allele). We calculated from those data the following parameters: allele frequency, total number of individuals, number of heterozygous individuals, and carrier frequency (CF). When calculating CF for a specific mutation or gene, only individuals who were heterozygous for a single mutation in the studied gene were considered, while homozygous individuals were excluded. However, it is important to note that this CF value may include individuals who are biallelic for mutations in a different AR-IEM gene that results in the same phenotype as the gene being studied.

### Hardy-Weinberg Equation

From those data, we want to know disease specific and pan-IEM disease prevalence and total number of carriers in the whole South Asian population. For these we will use the Hardy-Weinberg equation.

The Hardy-Weinberg equation (Hardy, 1908) is a fundamental principle in population genetics that describes how allele and genotype frequencies remain constant from generation to generation in an ideal non-evolving population by taking five assumptions about the population. The assumptions are: no new mutation is created, individuals in the population mate randomly, no natural selection resulting all individuals having an equal chance of survival and reproduction, the population is extremely large and there is no migration in or out of the population. Though our population does not ideally obey all the assumptions, but using the equation will give a pretty good assumption of our results. It states that if there are two alleles p & q for a particular position where p & q being frequency of the dominant allele and recessive allele respectively, and if sum of the two alleles frequency is one, i.e., p + q = 1, then: p^2^ + 2pq + q^2^ = 1. where the followings will be represented by each of the variable, p^2^ = frequency of the homozygous dominant genotype i.e., frequency of people who are neither carrier of the disease nor have the disease; 2pq = frequency of the heterozygous genotype i.e., frequency of healthy carries of the disease; q^2^ = frequency of the homozygous recessive genotype i.e., frequency of people with the disease

## RESULTS

### AR-IEM Gene Detection

The list of rare inborn errors of metabolism (ORPHA ID 68637) was extracted from Orphanet database in February 2020 which yielded a list of 883 disorders comprising of thirteen sub groups (Congenital disorder of glycosylation ORPHA:137; Disorder of amino acid and other organic acid metabolism ORPHA:79062; Disorder of biogenic amine metabolism and transport ORPHA:79214; Disorder of carbohydrate metabolism ORPHA:79161; Disorder of energy metabolism ORPHA:79200; Disorder of lipid metabolism ORPHA:309005; Disorder of lysosomal-related organelles ORPHA:309340; Disorder of metabolite absorption and transport ORPHA:309824; Disorder of porphyrin and heme metabolism ORPHA:309813; Disorder of purine or pyrimidine metabolism ORPHA:79224; Lysosomal disease ORPHA:68366; Other metabolic disease ORPHA:91088 and Peroxisomal disease ORPHA:68373). OMIM search for these diseases yielded 200 Autosomal Recessive Disorders associated with 235 genes (table 1).

**Table 1.**
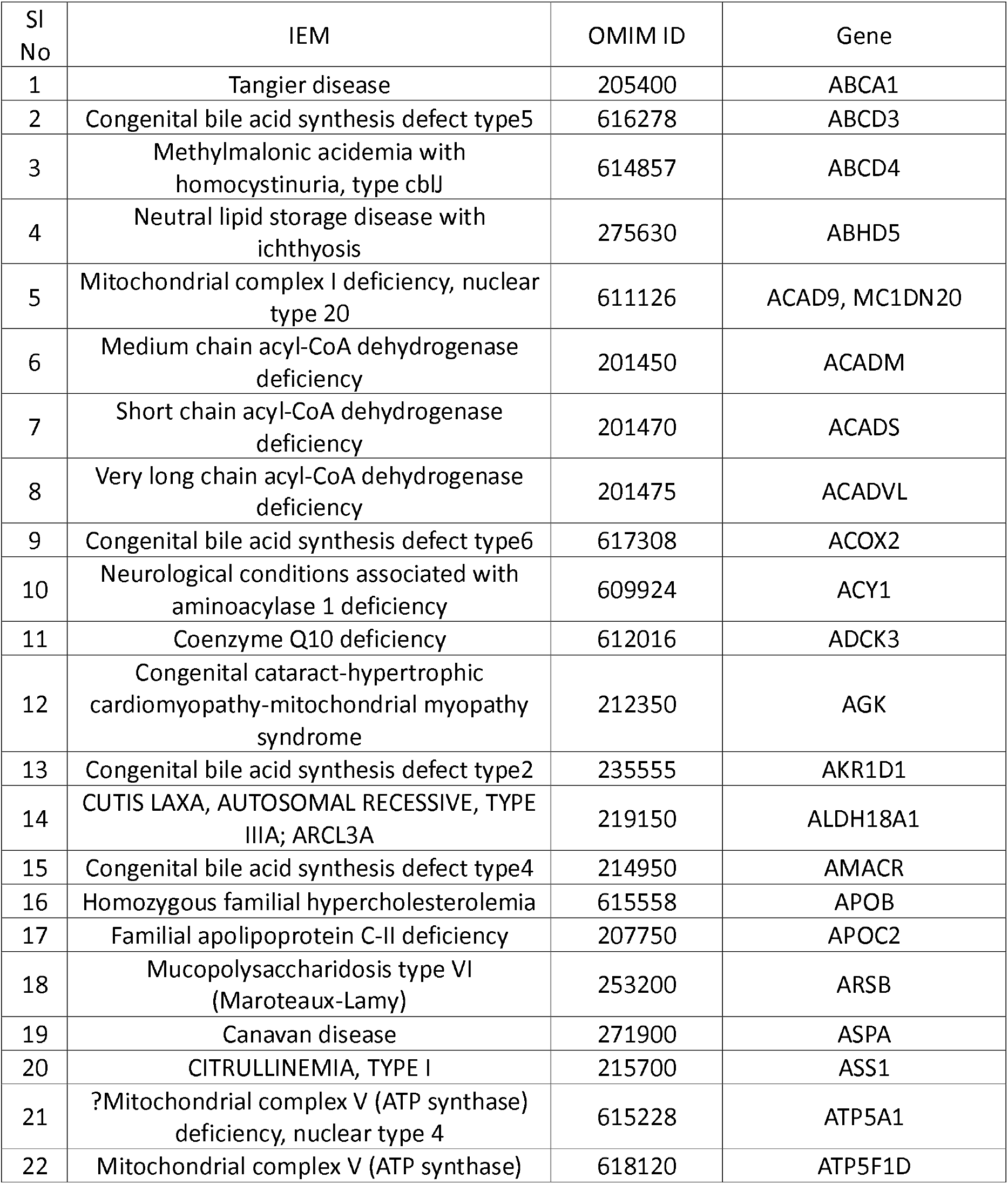
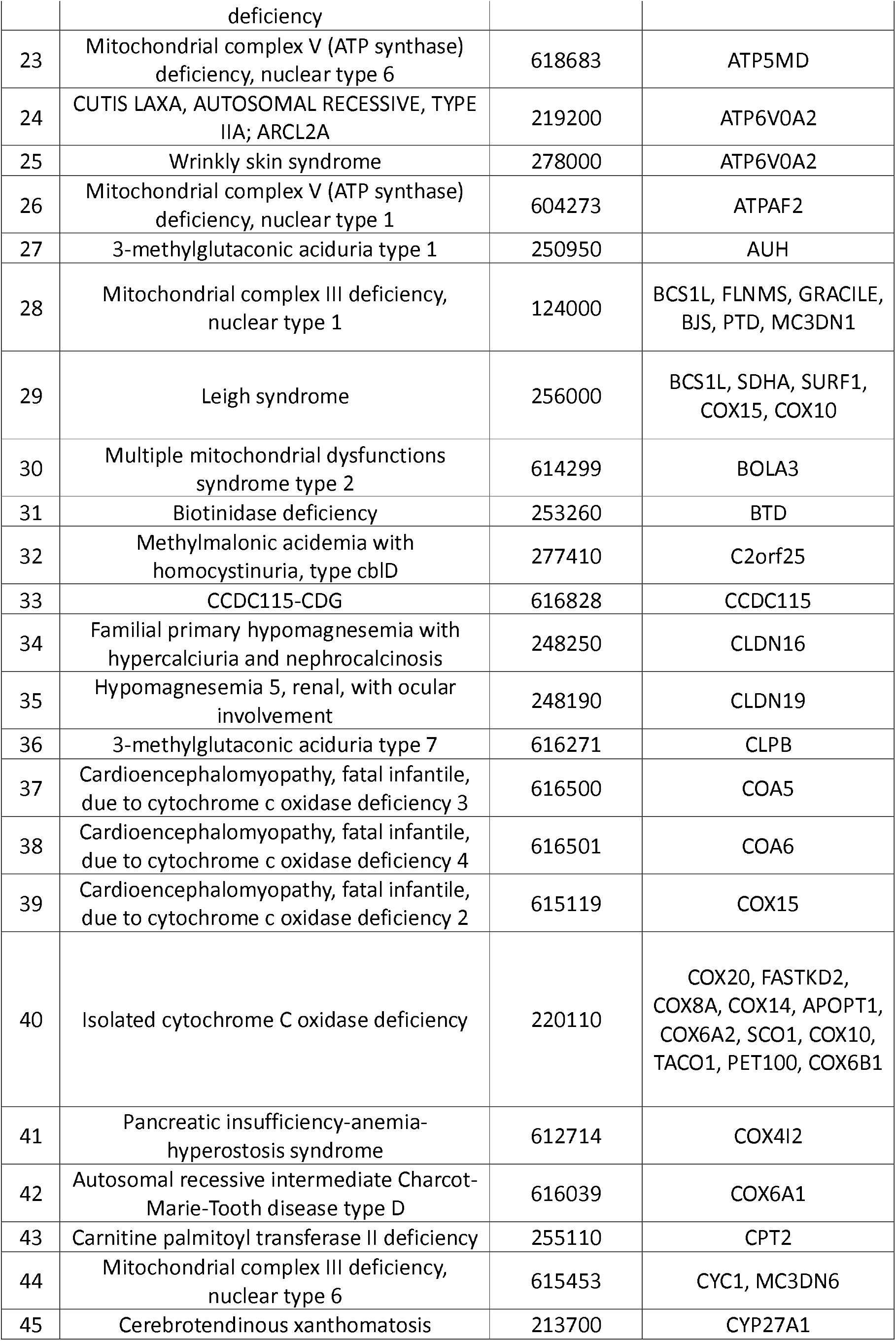
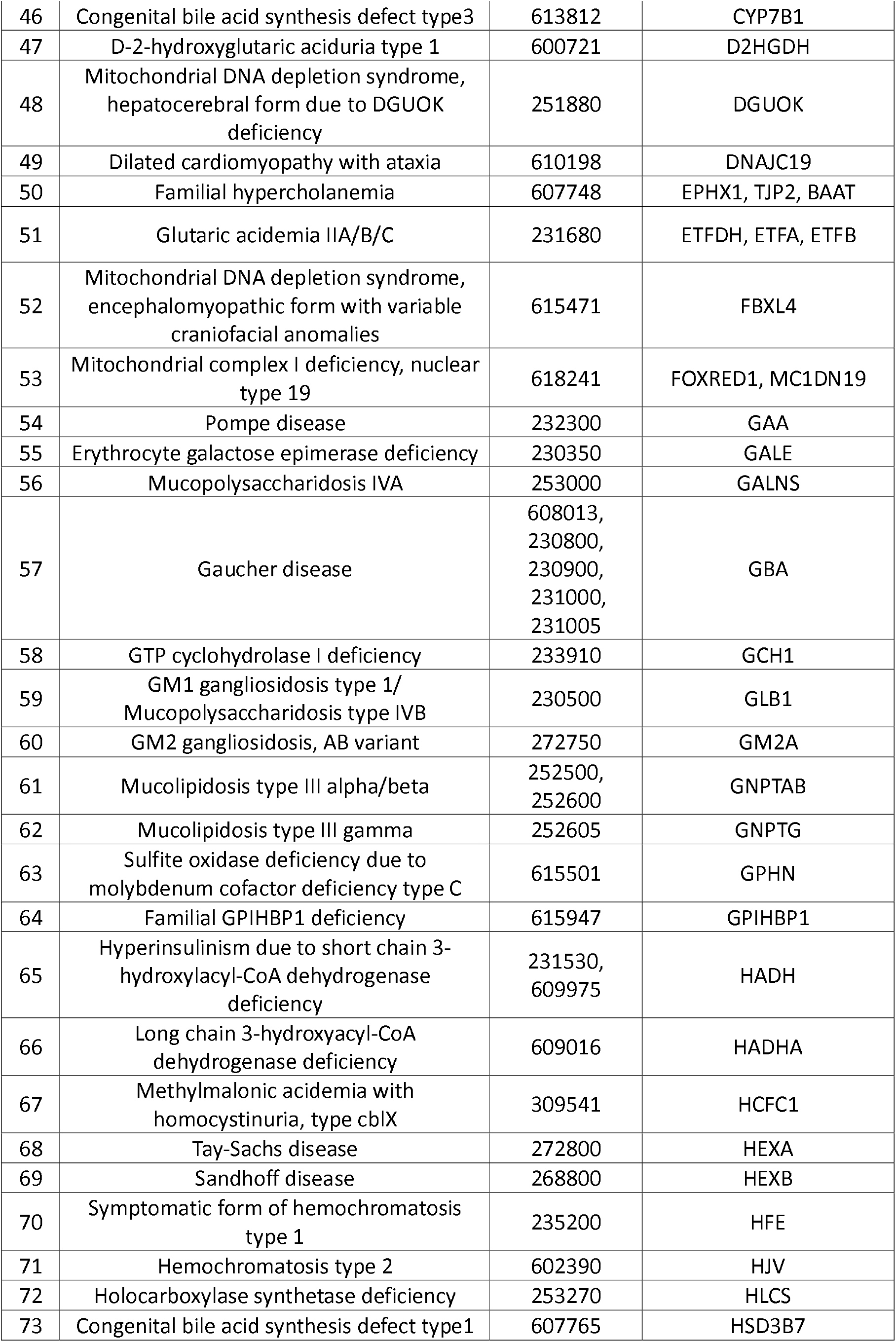
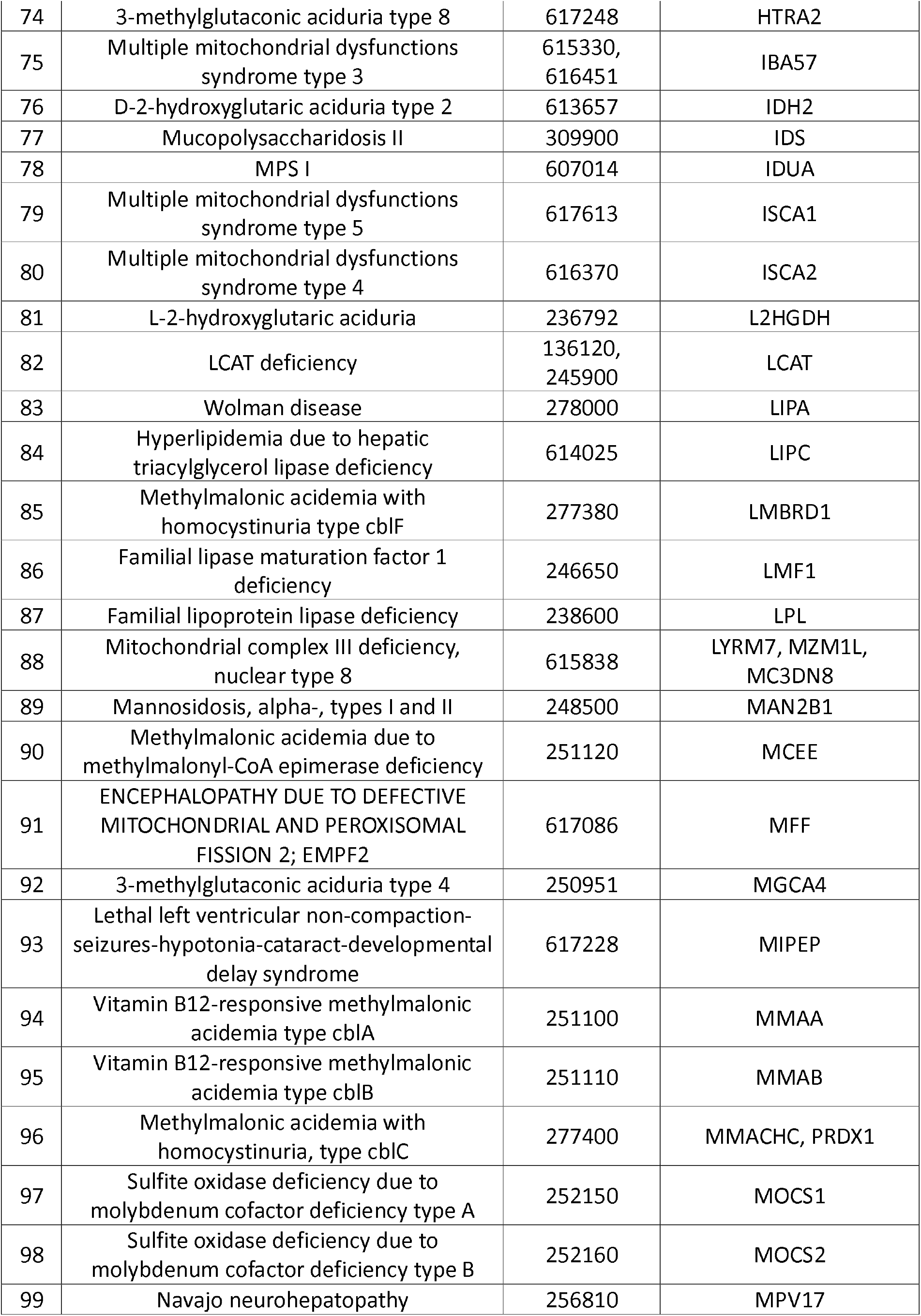
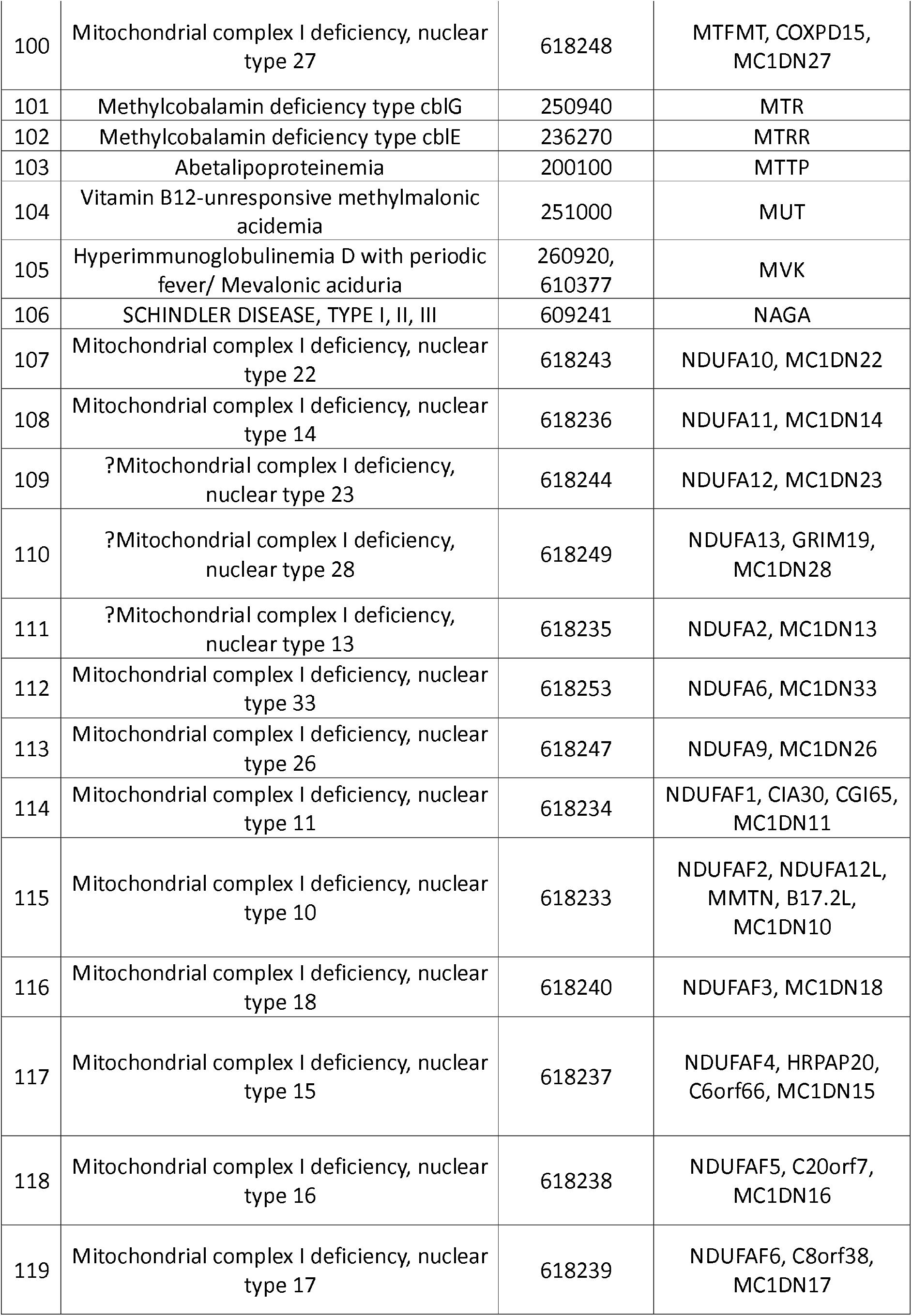
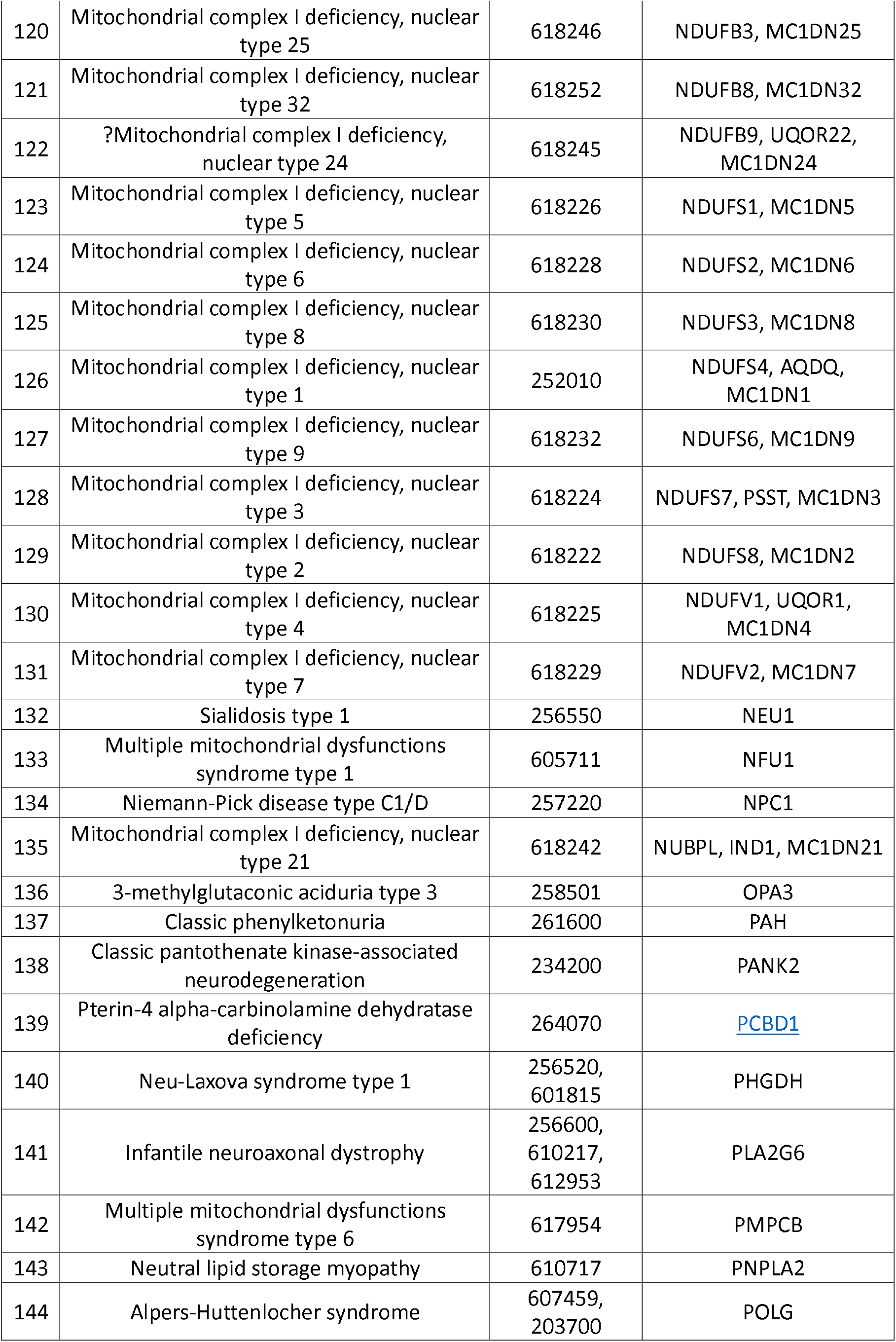
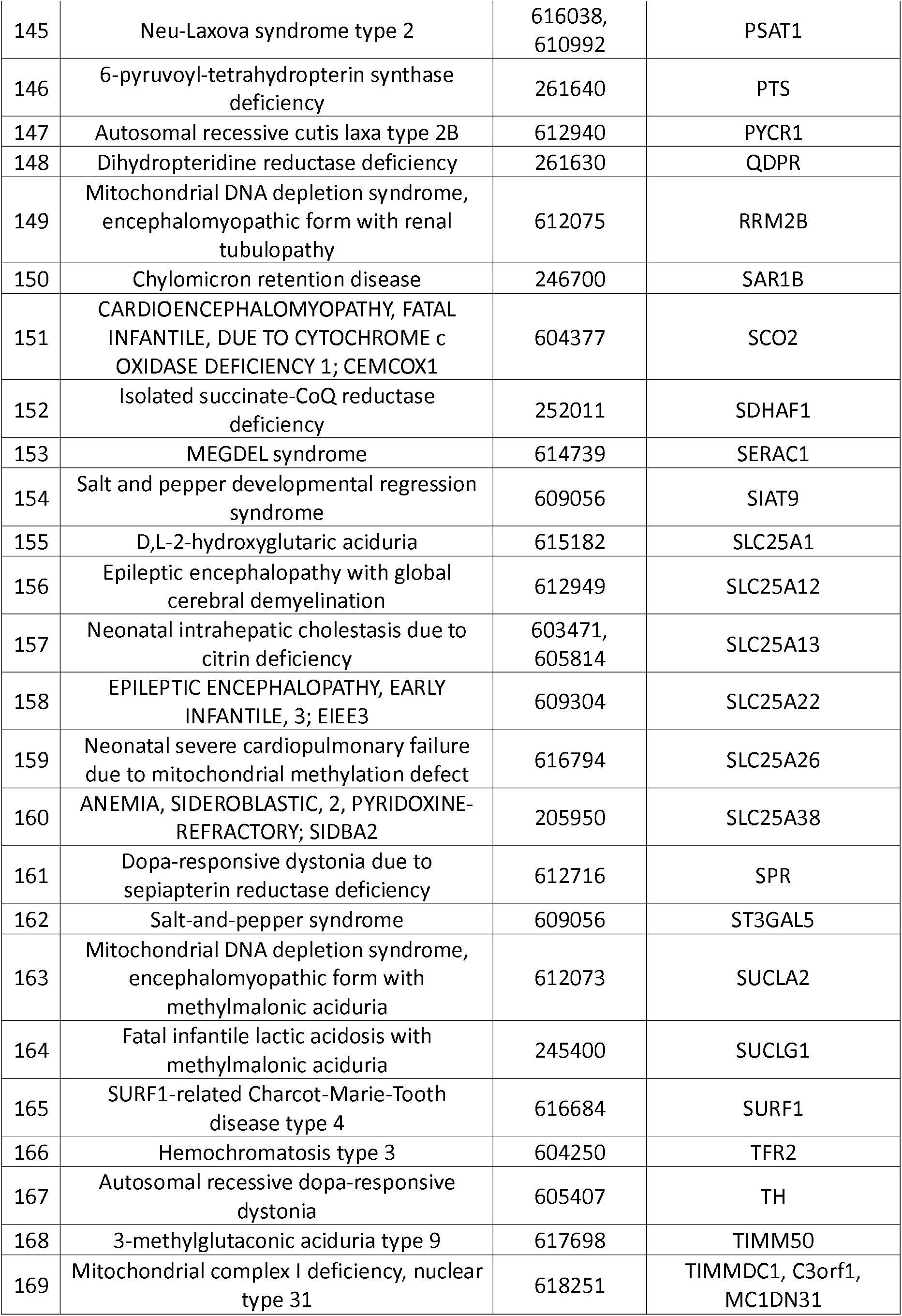
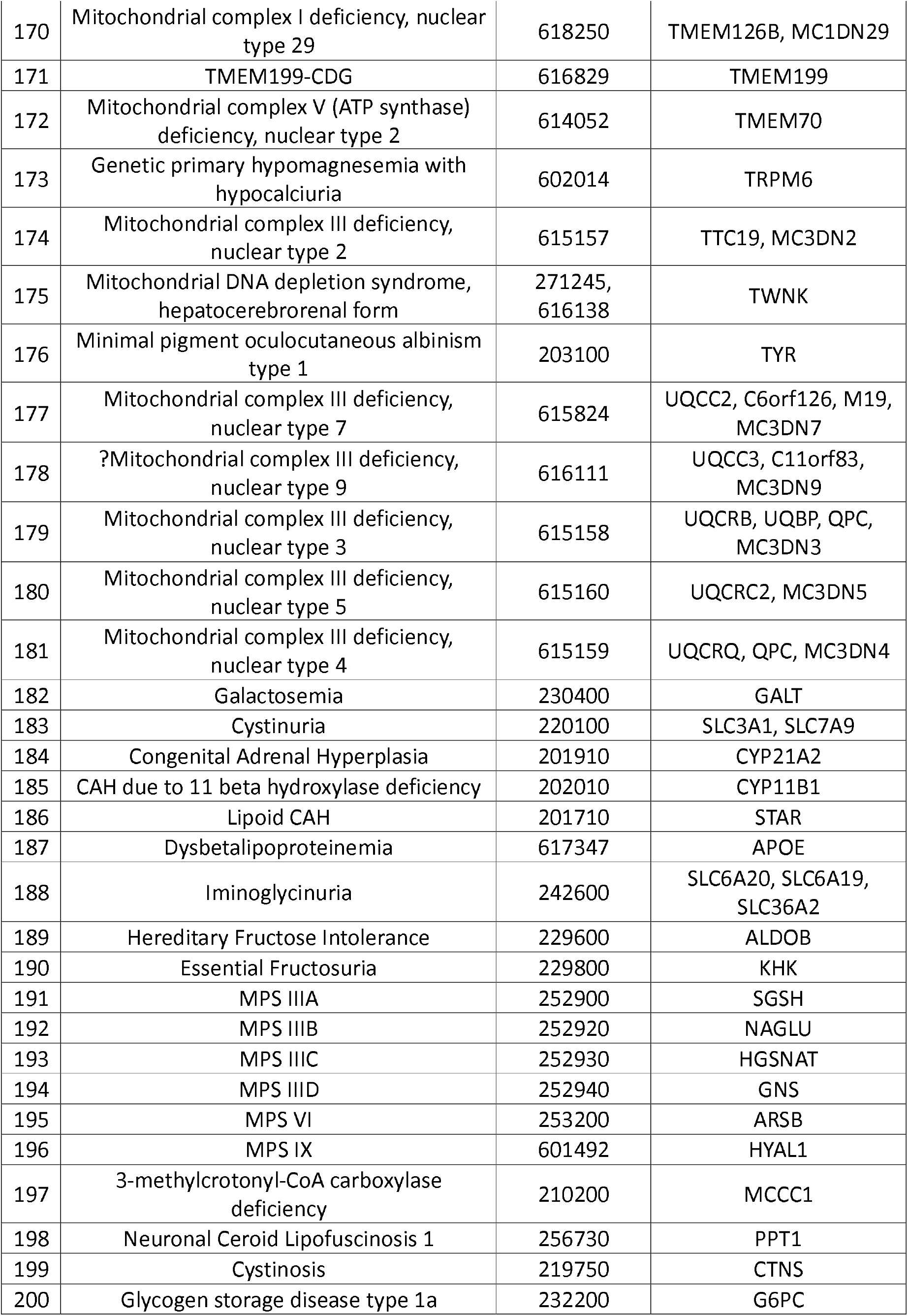
Autosomal Recessive Inborn Errors of Metabolism.

### Analysing the databases

After creating the three SQL databases from the gnomAD version 2.1 (https://gnomad.broadinstitute.org/), ClinVar (https://www.ncbi.nlm.nih.gov/clinvar/) and InterVar (https://wintervar.wglab.org/) database of the 221 AR-IEM genes, we started to detect total number of pathogenic DNA variants as per our methodology previously discussed.

A total number of 1,72,963 DNA variants were found from the gnomAD v2.1 dataset for the 221 AR-IEM genes. Among these 134 variants were found to be present only in one homozygous individual and thus excluded as they likely to represent nonreliable reads. The remaining 1,72,963 variants were divided in two subgroups: one with 7,200 probable truncating variants and 1,65,629 “other” variants.

We then validated the categorization information regarding 7,200 probable truncating variants as previously described methodology. Among them 6,308 variants were found to have both high-quality sequencing (as per gnomAD database) and allele frequency of less than or equal to 0.005; thus, were considered to be pathogenic. The rest 892 variants were searched in ClinVar and 167 were found to be reported as non-pathogenic. The remaining 725 variants were excluded as 715 had low-quality sequencing reported in gnomAD and 10 had allele frequency of greater than 0.005.

For the “other” 1,65,629 variants group, they were first searched in the SQL database of ClinVar and among 19,324 variants found in ClinVar 1,435 were labelled as “likely pathogenic” or “pathogenic” and thus were taken as pathogenic variants. The 1,46,305 variants that were not found in ClinVar were searched in InterVar. There among those found 324 were interpreted as “likely pathogenic” or “pathogenic” and thus taken as pathogenic variants.

Thus, a total of 8,067 variants were taken as pathogenic variants for the 221 AR-IEM genes for further analysis.

### Population Data

Demographic data on the worldwide populations (in terms of the number of individuals per each subpopulation) are based on the Department of Economic and Social Affairs of the United Nations Secretariat 2017 Revision published on June 2017 (https://www.un.org/development/desa/publications/world-population-prospects-the-2017-revision.html) as follows: total worldwide population size, 7.6 billion; Africans, 1,258,311,831; East Asians, 1,646,834,964; South Asians, 1,870,824,410; Finnish, 5,516,175; European (nonFinnish), 735,802,007; Latino, 646,059,776; and other populations that are not represented in the gnomAD database (Oceania, 40,722,400; Northern America, 360,963,0043; West Asia, 268,093,436; Southeastern Asia, 649,200,488; and South-Central Asia, 1,941,780,322).

### Calculation of Carrier Frequency (CF)

From the gnomAD database, we calculated the following parameters: allele frequency, total number of individuals, number of heterozygous individuals, and carrier frequency (CF). The combined minor allele frequency of the twenty most common IEMs for south Asian population are shown in the bar chart in Figure 2.

**Figure 1.**
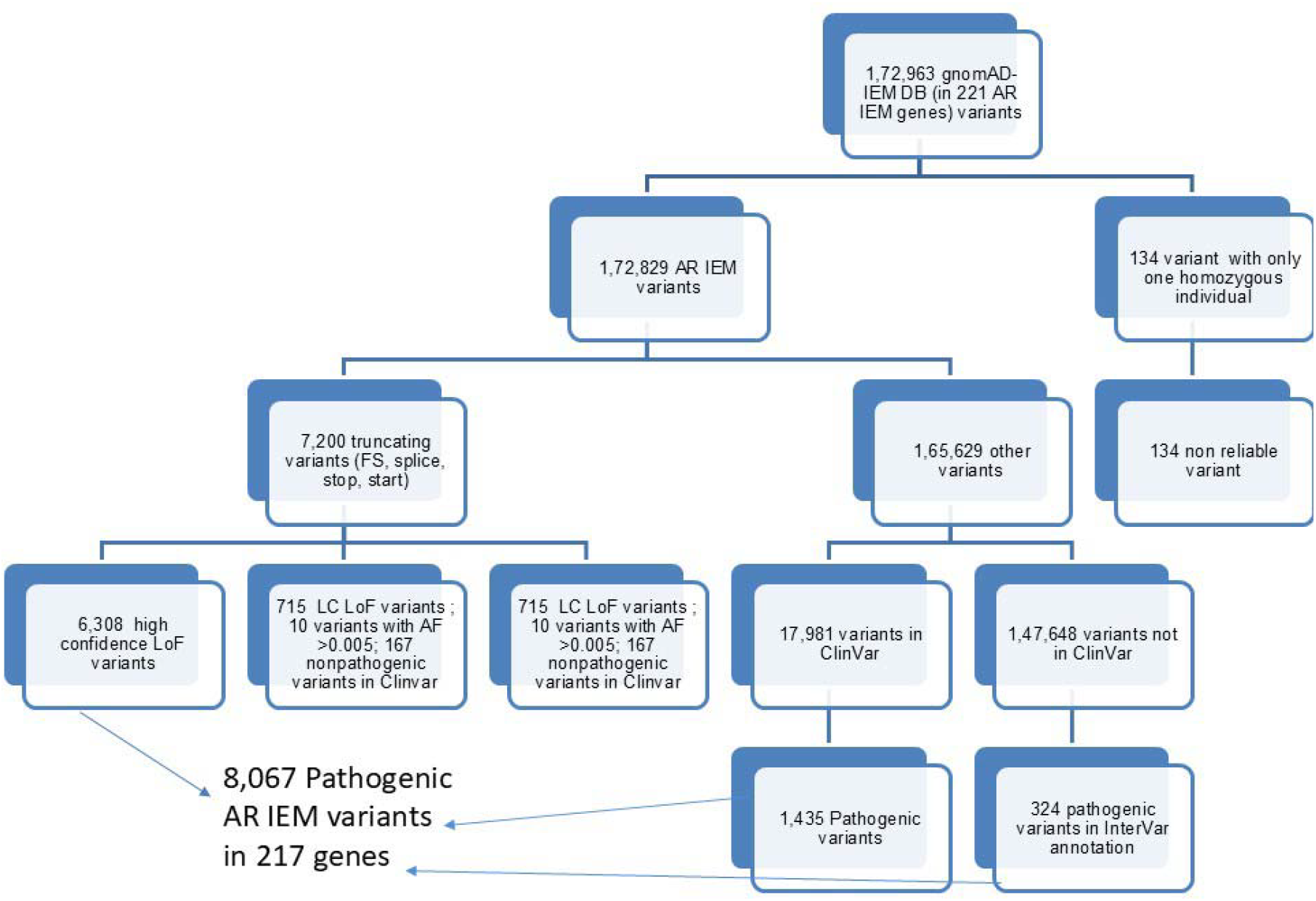

**Figure 2.**
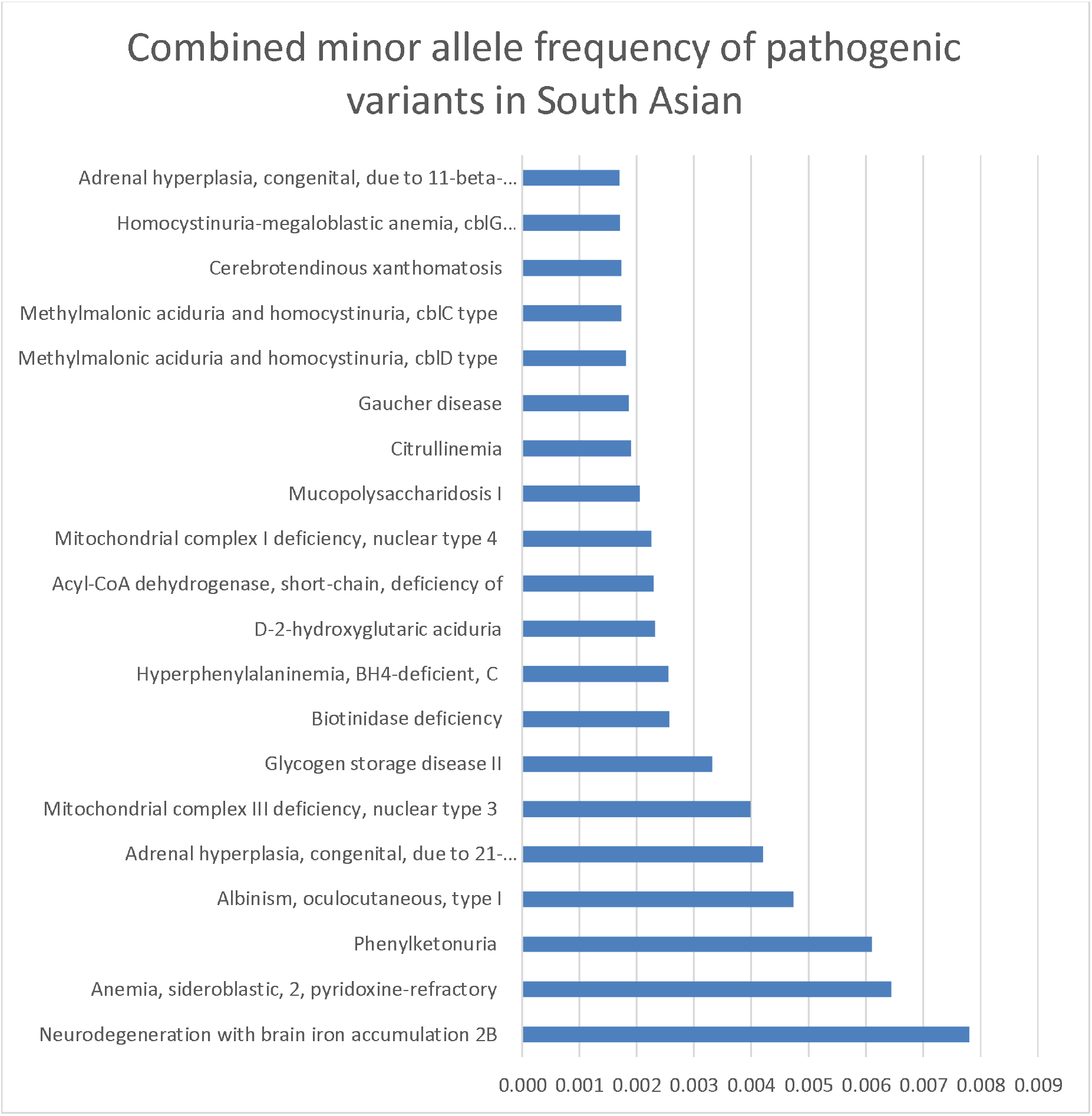

### Hardy-Weinberg Equation

Applying the equation, frequency of healthy carries of the disease (2pq) and frequency of people with the disease (q^2^) were calculated for all the subpopulations. Multiplying the frequency data with the total population data, the total number of estimates for both carriers and people with disease were calculated. These data have been shown in Figure 3 & 4.

**Figure 3.**
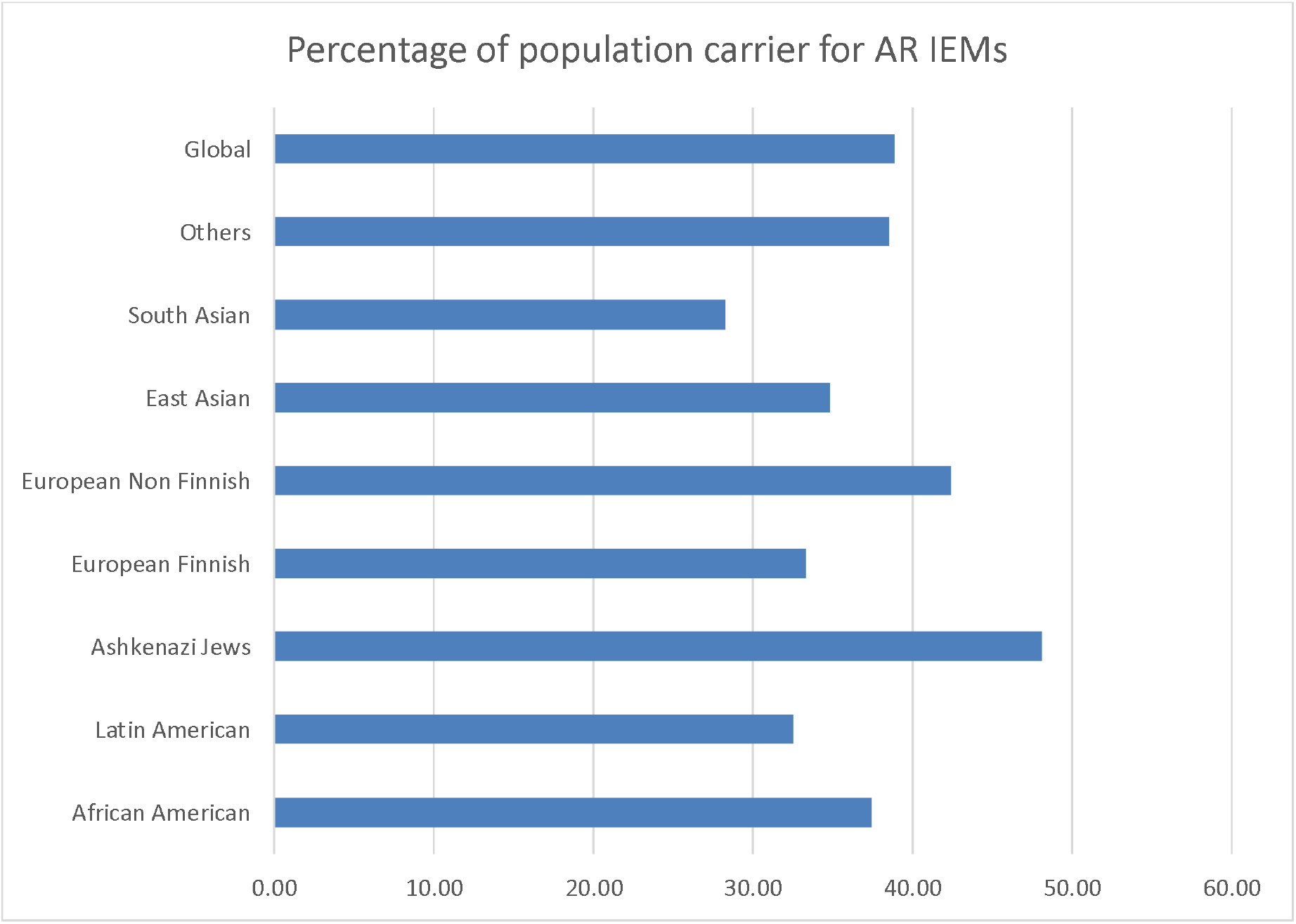

**Figure 4.**
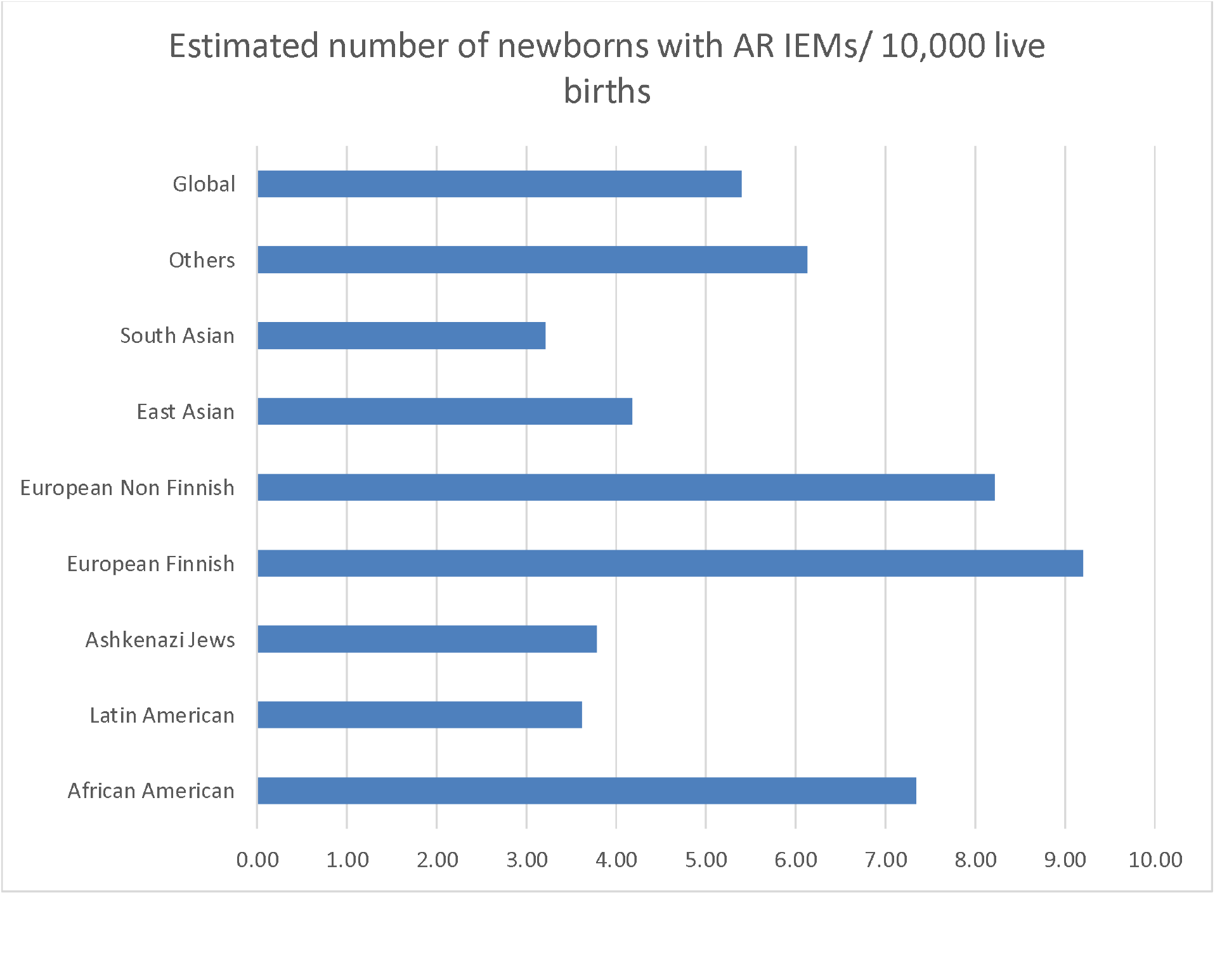

## DISCUSSION

As per our estimation almost one third of the Global population is carrier for a pathogenic variant responsible for rare autosomal recessive inborn error of metabolism with the highest carrier frequency in the Ashkenazi Jews. Globally per thousand live births approximately five children are born with an ARIEM. European Finnish have the highest burden of nine out of 10,000 live births. With 25 million live births per year India is expected to have at least 8,025 newborns with an ARIEM. Since many of these diseases are treatable (Figure 2) early newborn screening holds the key to ensure optimal management of these children.

There are several limitations in our study including the lack of corrections for unaffected carriers having have a heterozygous pathogenic mutation in multiple genes which needed correction by inclusion-exclusion principle, and the independence probability theory. Other limitations of this approach include presence of pseudogenes which might make NGS data unreliable. Also, we did not look for population frequency of copy number variations in ARIEM genes. Racial admixture cannot be ruled out in the gnomAD data. Hence, availability of Whole Genome data from representative healthy populations such as the Genome India data can help to refine these estimates.

## Data Availability

All data produced in the present study are available upon reasonable request to the authors

## REFERENCES

Ferreira, C. R., & van Karnebeek, C. D. M. (2019). Inborn errors of metabolism. Handbook of clinical neurology, 162, 449–481. 10.1016/B978-0-444-64029-1.00022-9

Krishnamurthy, J., Madhivanan, S., Kumarasamy, K., & Karthick, A. (2020). Clinical spectrum of inborn errors of metabolism in children in a tertiary care hospital. International Journal of Contemporary Pediatrics, 7(3), 495. 10.18203/2349-3291.ijcp20200500

Kruszka, P., & Regier, D. (2019). Inborn Errors of Metabolism: From Preconception to Adulthood. American family physician, 99(1), 25–32.

Lander, E. S., Linton, L., Birren, B. W., Nusbaum, C., Zody, M. C., Baldwin, J. N., … & Morgan, M. J. (2001). Initial sequencing and analysis of the human genome. Nature, 409(6822), 860–921. 10.1038/35057062

Lewis, C. and Vassos, E. (2020). Polygenic risk scores: from research tools to clinical instruments. Genome Medicine, 12(1). 10.1186/s13073-020-00742-5

Marian A. J. (2020). Clinical Interpretation and Management of Genetic Variants. JACC. Basic to translational science, 5(10), 1029–1042. 10.1016/j.jacbts.2020.05.013

Tabor, H. K., Auer, P. L., Jamal, S. M., Chong, J. X., Yu, J. H., Gordon, A. S., … & Bamshad, M. J. (2014). Pathogenic variants for Mendelian and complex traits in exomes of 6,517 European and African Americans: implications for the return of incidental results. American journal of human genetics, 95(2), 183–193. 10.1016/j.ajhg.2014.07.006

Therrell, B. L., Padilla, C. D., Borrajo, G. J. C., Khneisser, I., Schielen, P. C. J. I., Knight-Madden, J., Malherbe, H. L., & Kase, M. (2024). Current Status of Newborn Bloodspot Screening Worldwide 2024: A Comprehensive Review of Recent Activities (2020-2023). International journal of neonatal screening, 10(2), 38. 10.3390/ijns10020038

Vankwani, S., Wasim, M., Mirza, M. R., & Awan, F. R. (2024). Closing the gap: An urgent need for newborn screening of organic acid disorders in developing countries. JPMA. The Journal of the Pakistan Medical Association, 74(6), 1136–1143. 10.47391/JPMA.10223

Venter, J. C., Adams, M. D., Myers, E. W., Li, P. W., Mural, R. J., Sutton, G. G., … & Zhu, X. (2001). The sequence of the human genome. Science (New York, N.Y.), 291(5507), 1304– 1351. 10.1126/science.1058040

Hardy G. H. (1908). MENDELIAN PROPORTIONS IN A MIXED POPULATION. Science (New York, N.Y.), 28(706), 49–50. 10.1126/science.28.706.49

Richards, S., Aziz, N., Bale, S., Bick, D., Das, S., Gastier-Foster, J., … & ACMG Laboratory Quality Assurance Committee (2015). Standards and guidelines for the interpretation of sequence variants: a joint consensus recommendation of the American College of Medical Genetics and Genomics and the Association for Molecular Pathology. Genetics in medicine: official journal of the American College of Medical Genetics, 17(5), 405–424. 10.1038/gim.2015.30

